# Sex-specific multimorbidity clusters and all-cause mortality in relatively healthy older adults: findings from the ASPREE cohort

**DOI:** 10.64898/2026.06.18.26355909

**Authors:** Rong Du, Swarna Vishwanath, Zimu Wu, Aung Zaw Zaw Phyo, Suzanne G. Orchard, Kerry Sheets, Michael E. Ernst, Joanne Ryan

**Author notes:** **Corresponding Author:** Professor Joanne Ryan, School of Public Health and Preventive Medicine, Monash University, 553 St Kilda Road, Melbourne, VIC 3004, Australia.

## Abstract

**Background:** Multimorbidity is common in older adults, but sex differences in chronic condition clustering remain unclear. This study explored multimorbidity clusters and their associations with all-cause mortality among community-dwelling adults aged 70 years and over.

**Methods:** This was a secondary analysis of data from 16,095 Australian ASPREE participants aged ≥70 years without prior dementia or cardiovascular disease. Fifteen baseline chronic conditions were grouped using latent class analysis (LCA). Observed-to-expected (O/E) ratios characterised conditions over-represented within clusters, and Cox proportional hazards models assessed associations with all-cause mortality.

**Results:** Among 16,095 participants (mean age 74 years), 88.3% had multimorbidity at baseline; 4,217 deaths occurred over a median follow-up of 10.85 years. Five clusters were identified overall: hypertension & dyslipidemia (52.1%), gout & metabolic (14.4%), depressive symptoms, osteoporosis & frailty (10.0%), anaemia & kidney disease (10.2%), and hypotension, thyroid disorder & past cancer (13.3%). Sex-stratified analyses revealed three clusters in males and four in females. The frailty, depressive symptoms & osteoporosis cluster was associated with higher mortality in both sexes (aHR 1.56 [95% CI 1.40–1.73] in males; 1.68 [1.49–1.89] in females). Higher mortality was also observed for the metabolic, gout & kidney disease cluster in males (aHR 1.63 [1.47–1.81]) and the gout, anaemia & kidney disease cluster in females (aHR 1.96 [1.74–2.21]).

**Conclusions:** Distinct multimorbidity clusters differed by sex and were associated with increased all-cause mortality. These findings may support risk stratification, targeted screening, and more person-centred management of older adults with multimorbidity.

## Introduction

Multimorbidity, commonly defined as the coexistence of two or more chronic conditions, is becoming increasingly prevalent with the aging population, posing a major challenge for public health and healthcare systems^1,2^. The prevalence of multimorbidity rises sharply with age, affecting approximately 65% of adults aged 65 years or older and 80% of those aged 85 years or older(2). This growing burden is associated with poorer quality of life, functional decline, psychological distress, greater healthcare use, polypharmacy, and increased mortality(3). However, its management remains complex because clinical guidelines and healthcare delivery models often focus on single diseases, making it difficult to address the needs of older adults with diverse combinations of coexisting chronic conditions(3,4).

Multimorbidity is often measured using disease count, providing an intuitive summary of overall disease burden(5). Beyond disease count, cluster-based approaches can clarify the non-random co-occurrence of chronic conditions and identify specific disease combinations with distinct prognostic implications(6–8). These clusters can be derived using predefined clinical groupings, frequently co-occurring conditions, or data-driven approaches(7,9), offering clinically relevant information with treatment implications beyond simple disease counts by identifying subgroups with distinct risk profiles(10,11).

Existing studies of multimorbidity clusters show substantial heterogeneity(8,12,13). Nevertheless, large systematic reviews have consistently identified cardiometabolic or cardiovascular-related clusters, often mixing chronic conditions such as hypertension and diabetes, with major health events like ischaemic heart disease, heart failure, and stroke(8,12). Less is known about multimorbidity clustering in community-dwelling older adults without major clinical cardiovascular disease, as well as possible sex differences(14–16). Moreover, among the few sex-stratified analyses conducted to date, most study populations have been from Europe, Asia, and North America, and the largely cross-sectional design of these studies has limited their ability to examine the longer-term prognostic implications of multimorbidity clusters(8).

Using data from a large cohort of relatively healthy community-dwelling adults aged 70 years or older in Australia, this study aimed to characterise sex-specific multimorbidity clusters and their association with all-cause mortality.

## Methods

### Study population

Data came from the Aspirin in Reducing Events in the Elderly (ASPREE) trial and its ongoing observational extension study (ASPREE-XT). ASPREE was a double-blind, randomised, placebo-controlled trial that examined the effects of daily 100 mg aspirin on various health outcomes in older adults(17,18). The present analysis included Australian ASPREE participants, who were recruited between March 2010 and December 2014 through their usual general practitioners, and were community-dwelling adults aged 70 years or older, free from prior cardiovascular disease, dementia, or major physical disability at trial entry(19). At baseline and annual study visits, trained staff conducted in-person interviews to collect information on participant characteristics, health status, and medical history, followed by laboratory assessments and review of medication use. Baseline covariates included age, sex, education, residence area, living status, smoking status, and alcohol consumption.

### Assessment of chronic conditions

At baseline, 22 chronic conditions were initially identified in the ASPREE cohort using direct physical and laboratory measurements, self-report, and/or concomitant medication use, with thresholds defined according to clinical guidelines(19,20). For the present analysis, conditions were selected based on clinical relevance, data completeness, and sufficient prevalence to support stable latent class analysis. Seven conditions were excluded: Parkinson’s disease and urinary incontinence due to their prevalence being below 2%; while pneumonia, deep vein thrombosis, macular degeneration, gastro-oesophageal reflux, and osteoarthritis were excluded as they were only captured on a subsample of the population (with >50% missing data). The final analysis included 15 chronic conditions: hypertension, hypotension, diabetes, dyslipidemia, obesity, thyroid disorder, obstructive pulmonary disease (OPD), kidney disease, frailty, cancer history, bowel polyp, gout, osteoporosis, anaemia, and depressive symptoms. Definitions and data sources for each condition are provided in **eTable 1**. All conditions were ascertained at enrolment and were considered to reflect participants’ baseline health status.

### All-cause mortality

All-cause mortality was defined as death from any cause(21). Deaths were ascertained through medical records, reports from next of kin or close contacts during routine follow-up, and linkage to the Australian National Death Index. Follow-up time was calculated from recruitment to death or censoring, defined as loss to follow-up or the last study visit, whichever occurred first. For the current analysis, deaths up to the end of December 2024 were included in the latest release of the ASPREE-XT06 dataset.

### Statistical analysis

#### Latent class analysis

Latent class analysis (LCA)(22) was used to identify multimorbidity clusters based on the co-occurrence of chronic conditions in the overall sample and then separately in males and females. LCA is a person-centered probabilistic approach that classifies individuals into mutually exclusive latent groups according to similar response clusters(22). In the overall sample, models with two to ten classes were evaluated, whereas separate models with two to eight classes were assessed in males and females. This range was selected to allow sufficient exploration of potential class solutions while avoiding overly complex or unstable models with small class sizes, particularly in the sex-stratified analyses. The optimal number of classes was selected by considering model fit indices, including Akaike’s Information Criterion (AIC), Bayesian Information Criterion (BIC), log-likelihood, and the likelihood ratio chi-square statistic (G²), as well as class size and clinical interpretability(11). To avoid unstable or poorly informative solutions, each class was required to include at least 10% of the study population.

After class selection, the identified latent classes were further characterised by the distribution of chronic conditions within each cluster. For each condition, we calculated the observed probability within each cluster and the expected probability in the corresponding analytical sample. The observed-to-expected (O/E) ratio was then derived to quantify relative over-representation, with values greater than one indicating that a condition was more common within a given cluster than expected based on its prevalence in the corresponding sample.

Cluster labels were assigned primarily according to the two or three conditions with the highest O/E ratios within each cluster, reflecting conditions that were disproportionately represented. When the same condition was overrepresented across multiple clusters, it was included in the label only for the cluster with the highest O/E ratio.

### Survival analysis

Kaplan–Meier survival curves were generated to describe survival according to multimorbidity clusters in the overall sample and in sex-stratified analyses, with between-cluster differences assessed using the log-rank test. Mortality rates were calculated as the number of deaths divided by total person-years of follow-up and expressed per 1,000 person-years with 95% confidence intervals (CIs). Cox proportional hazards models were used to estimate the associations between membership in multimorbidity clusters at baseline and all-cause mortality in the corresponding analytical samples. The proportional hazards assumption was assessed using Schoenfeld residuals, and the main exposure variable satisfied the assumption. Results are presented as hazard ratios (HRs) with 95% CIs. For each analysis, the reference cluster was selected based on two criteria: the lowest relative over-representation of chronic conditions in the corresponding analytical sample, assessed using the O/E ratio distribution and clinical interpretability, and the lowest observed mortality incidence. Three models were fitted. Model 1 was adjusted for age and sex in the overall analysis. Model 2 was additionally adjusted for education, residence area, and living status. Model 3 was further adjusted for smoking status and alcohol consumption.

### Sensitivity analyses

Sensitivity analyses were conducted to assess the robustness of the cluster–mortality associations. First, the individual-level number of chronic conditions was added to the fully adjusted Cox proportional hazards models to determine whether the associations reflected multimorbidity clusters rather than simply overall disease burden. Second, to account for possible changes in disease clusters over time, we examined the association between baseline cluster membership and 5-year all-cause mortality using the same fully adjusted Cox models as in the main analysis.

Baseline characteristics and chronic conditions were summarised as n (%) for categorical variables and mean (SD) for continuous variables. Between-group differences were assessed using chi-square tests or analysis of variance, as appropriate. Statistical analyses were conducted using R version 4.4.2 (R Foundation for Statistical Computing, Vienna, Austria), with the poLCA and survival packages. All P values were two-sided, and statistical significance was defined as P < 0.05.

## Results

### Participant characteristics and prevalence of chronic conditions

Among the 16,703 Australian ASPREE participants, 608 were excluded due to missing data on at least one of the 15 chronic conditions, leaving 16,095 participants in the final analytical sample. Of the included participants, 8,885 (55.2%) were female, and 7,210 (44.8%) were male. Compared with males, females were slightly older, more likely to live alone, and less likely to be current drinkers or former smokers (**Table 1**). Overall, 14,205 participants (88.3%) had multimorbidity, defined as the presence of two or more of the 15 chronic conditions. Multimorbidity was more common in females than in males (90.4% vs. 85.6%), and females had a higher mean number of chronic conditions (3.4 [SD 1.6] vs. 3.1 [SD 1.6]).

**Table 1.** Baseline characteristics of Australian ASPREE participants in the overall sample and by sex.

| Characteristics | Overall<br>(N = 16,095) | Male<br>(N = 7,210) | Female<br>(N = 8,885) | P value |
| --- | --- | --- | --- | --- |
| Age, mean (SD) | 75.3 (4.4) | 75.1 (4.3) | 75.4 (4.4) | <0.001 |
| Age group, n (%) |  |  |  | <0.001 |
| 70–74 | 9318 (57.9) | 4314 (59.8) | 5004 (56.3) |  |
| 75–79 | 4274 (26.6) | 1832 (25.4) | 2442 (27.5) |  |
| ≥80 | 2503 (15.6) | 1064 (14.8) | 1439 (16.2) |  |
| Education, n (%) |  |  |  | <0.001 |
| ≥12 years | 8006 (49.7) | 3818 (53.0) | 4188 (47.1) |  |
| <12 years | 8089 (50.3) | 3392 (47.0) | 4697 (52.9) |  |
| Residence area, n (%) |  |  |  | 0.749 |
| Outer region | 1886 (11.7) | 831 (11.5) | 1055 (11.9) |  |
| Inner region | 5743 (35.7) | 2588 (35.9) | 3155 (35.5) |  |
| Major cities | 8466 (52.6) | 3791 (52.6) | 4675 (52.6) |  |
| Living status, n (%) |  |  |  | <0.001 |
| Living with someone | 10969 (68.2) | 5761 (79.9) | 5208 (58.6) |  |
| Living alone at home | 5126 (31.8) | 1449 (20.1) | 3677 (41.4) |  |
| Smoking status, n (%) |  |  |  | <0.001 |
| Current | 544 (3.4) | 278 (3.9) | 266 (3.0) |  |
| Former | 6582 (40.9) | 3835 (53.2) | 2747 (30.9) |  |
| Never | 8969 (55.7) | 3097 (43.0) | 5872 (66.1) |  |
| Alcohol consumption, n (%) |  |  |  | <0.001 |
| Current | 12700 (78.9) | 6155 (85.4) | 6545 (73.7) |  |
| Former | 781 (4.9) | 421 (5.8) | 360 (4.1) |  |
| Never | 2614 (16.2) | 634 (8.8) | 1980 (22.3) |  |
| Chronic condition count, n (%) |  |  |  | <0.001 |
| 0 | 319 (2.0) | 204 (2.8) | 115 (1.3) |  |
| 1 | 1571 (9.8) | 831 (11.5) | 740 (8.3) |  |
| ≥2 | 14205 (88.3) | 6175 (85.6) | 8030 (90.4) |  |
| Chronic condition count,<br>mean (SD) | 3.3 (1.6) | 3.1 (1.6) | 3.4 (1.6) | <0.001 |

Note: P values compare males and females. Continuous variables were compared using t-tests, and categorical variables were compared using chi-square tests.

In the overall sample and in both sexes, hypertension, dyslipidemia, and frailty were the most common conditions (**Table 2**), with hypertension being slightly more prevalent in males (76.4% in males vs. 74.3% in females), while dyslipidemia (76.9% in females vs. 56.8% in males) and frailty (39.7% in females vs. 37.3% in males) were more prevalent in females. Among the other conditions, the most pronounced sex differences were observed for osteoporosis (9.0% in females vs. 1.9% in males), gout (1.2% in females vs. 7.8% in males), and depressive symptoms (11.4% in females vs. 7.4% in males).

**Table 2.** Prevalence of the 15 included chronic conditions in the overall sample and by sex.

| Chronic condition | Overall<br>(N = 16,095) | Male<br>(N = 7,210) | Female<br>(N = 8,885) | P value |
| --- | --- | --- | --- | --- |
| Hypertension | 12110 (75.2) | 5508 (76.4) | 6602 (74.3) | 0.002 |
| Hypotension | 542 (3.4) | 162 (2.3) | 380 (4.3) | <0.001 |
| Diabetes | 1587 (9.9) | 871 (12.1) | 716 (8.1) | <0.001 |
| Dyslipidemia | 10927 (67.9) | 4095 (56.8) | 6832 (76.9) | <0.001 |
| Obesity | 4634 (28.8) | 1869 (25.9) | 2765 (31.1) | <0.001 |
| Thyroid disorder | 1131 (7.0) | 166 (2.3) | 965 (10.9) | <0.001 |
| Obstructive pulmonary disease | 1950 (12.1) | 822 (11.4) | 1128 (12.7) | 0.013 |
| Kidney disease | 3351 (20.8) | 1454 (20.2) | 1897 (21.4) | 0.069 |
| Frailty | 6215 (38.6) | 2691 (37.3) | 3524 (39.7) | 0.003 |
| Cancer history | 3133 (19.5) | 1565 (21.7) | 1568 (17.7) | <0.001 |
| Bowel polyp | 3230 (20.1) | 1574 (21.8) | 1656 (18.6) | <0.001 |
| Gout | 670 (4.2) | 564 (7.8) | 106 (1.2) | <0.001 |
| Osteoporosis | 934 (5.8) | 136 (1.9) | 798 (9.0) | <0.001 |
| Anaemia | 653 (4.1) | 290 (4.0) | 363 (4.1) | 0.871 |
| Depressive symptoms | 1545 (9.6) | 535 (7.4) | 1010 (11.4) | <0.001 |

Note: Values are presented as n (%). P values compare males and females using chi-square tests.

### Multimorbidity clusters in the overall sample and by sex

Five multimorbidity clusters were identified in the overall sample **(Figure 1**). The largest cluster (n = 8,388, 52.1%) had the least distinctive chronic conditions profile, with only hypertension and dyslipidemia being slightly more common than expected (O/E ratio just above 1). The remaining clusters showed more distinct profiles of chronic conditions. The 2^nd^ most common cluster (n = 2,320, 14.4%) showed high representation of diabetes, gout, and obesity; while the 3^rd^ cluster (n = 1,616, 10.0%) was characterised by a high O/E ratio for depressive symptoms, together with osteoporosis, frailty, thyroid disease, and OPD. Anaemia and kidney disease clustered together in another cluster (n = 1,635, 10.2%), but the 5^th^ cluster, which had the greatest over-representation of hypotension (n = 2,136, 13.3%), showed more moderate representation for other conditions.

**Figure 1.**
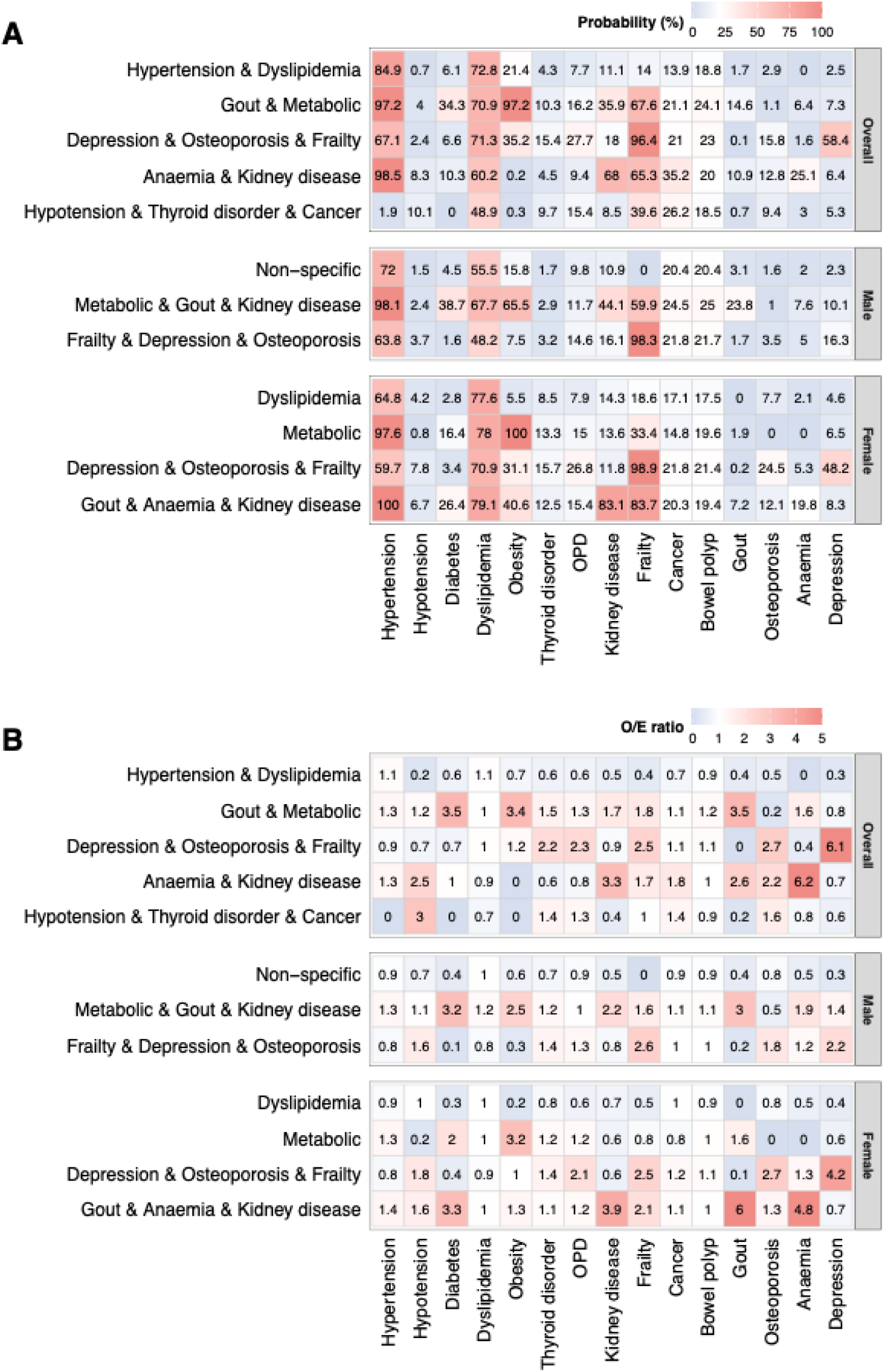
Baseline chronic condition profiles across multimorbidity clusters overall and by sex. (A) Conditional probabilities of 15 baseline chronic conditions within each cluster. (B) Observed-to-expected (O/E) ratios of chronic conditions within each cluster. “Metabolic” in cluster labels indicates obesity and diabetes. OPD = obstructive pulmonary disease. Alt text: Heatmaps comparing conditional probabilities and observed-to-expected ratios of baseline chronic conditions across multimorbidity clusters in the overall sample, males, and females.

The optimal number of clusters was three for males and four for females **(Figure 1)**. In males, one cluster was labelled non-specific because all chronic conditions had O/E ratios below 1 (n = 3,791, 52.6%), whereas in females, the comparable low-specificity cluster (n = 4,973, 56.0%) was mainly characterised by dyslipidaemia. Depressive symptoms, osteoporosis, and frailty clustered together in both sexes, but the female cluster showed a more pronounced profile, particularly for depressive symptoms(O/E ratio 4.2 vs 2.2), as well as higher over-representation of hypotension and thyroid disorder. The remaining male cluster had over-representation of diabetes (3.2), gout (3.1), obesity (2.5) and kidney disease (2.2), whereas females had two additional clusters: a metabolic cluster, mainly characterised by obesity (3.2) and to a lesser extent diabetes (2.0), and a cluster with high O/E ratios for gout (6.0), anaemia (4.8), and kidney disease (3.9), as well as diabetes (3.3).

Conditional probabilities of individual conditions within each cluster are shown in **Figure 1** and indicate that hypertension and dyslipidemia were common across several clusters, whereas conditions such as depressive symptoms, osteoporosis, gout, anaemia, kidney disease, and frailty showed more cluster-specific patterns.

### Baseline characteristics across multimorbidity clusters

Baseline characteristics varied significantly across multimorbidity clusters in the overall and sex-stratified analyses (**eTables 2-4**). Participants in the gout and metabolic cluster had the highest mean disease count and were more likely to have less than 12 years of education. In sex-stratified analyses, the frailty, depressive symptoms, and osteoporosis cluster in males had the oldest age profile, while in females, it was the gout, anaemia, and kidney disease cluster.

### Multimorbidity clusters and all-cause mortality

Over a median follow-up of 10.85 years (IQR: 9.63–12.08), 4,217 deaths occurred, including 2,160 in males and 2,057 in females, corresponding to a mortality rate of 29.35 per 1,000 person-years and 22.01 per 1,000 person-years, respectively (**eTable 5**). Kaplan–Meier survival curves showed that two clusters had significantly higher survival probability than the other three (log-rank test, P < 0.001) (**Figure 2**).

**Figure 2.**
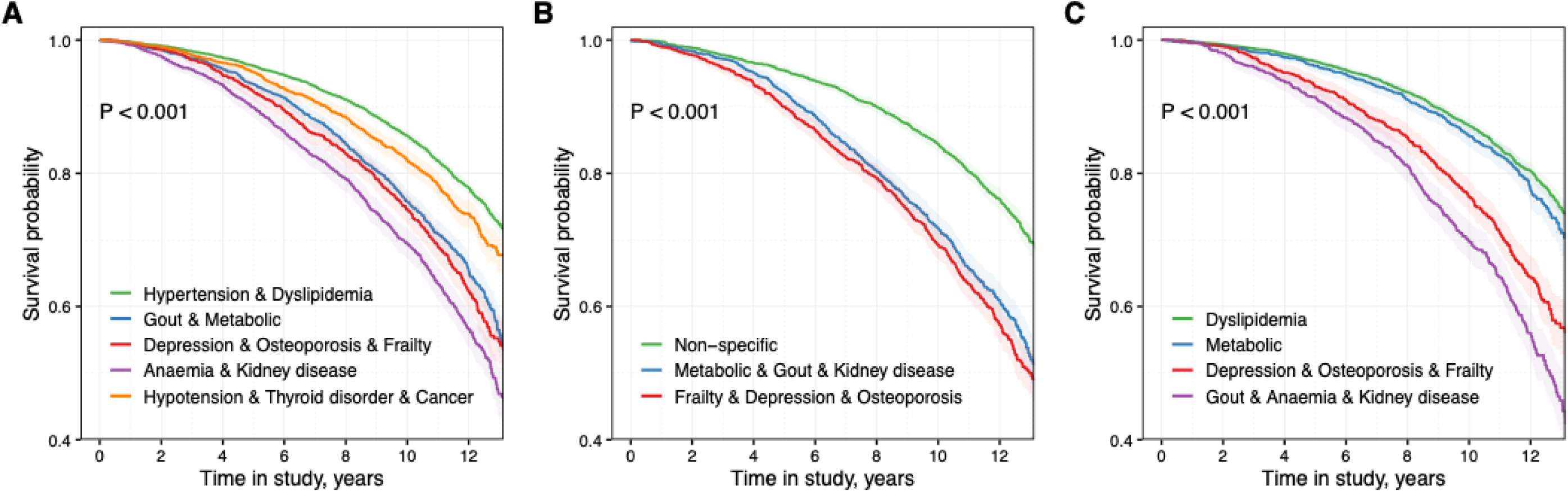
Kaplan–Meier survival curves by baseline multimorbidity cluster in the overall sample (A), males (B), and females (C). “Metabolic” in cluster labels indicates obesity and diabetes. Alt text: Kaplan–Meier survival curves showing differences in survival probability across baseline multimorbidity clusters in the overall sample, males, and females.

In the overall sample, all other clusters showed increased all-cause mortality compared with the hypertension and dyslipidemia cluster. The effect size was similar for the depressive symptoms, osteoporosis, and frailty cluster (aHR 1.68, 95% CI 1.52–1.85), the anaemia and kidney disease cluster (aHR 1.65, 95% CI 1.51–1.81), and the gout and metabolic cluster (aHR 1.61, 95% CI 1.47–1.75). The hypotension, thyroid disease, and past cancer cluster showed a more modest association with mortality (aHR 1.19, 95% CI 1.08–1.31) **(Figure 3)**. In sex-specific Cox models, relative to the non-specific cluster in males and the dyslipidaemia cluster in females, stronger mortality associations were observed for the metabolic, gout, and kidney disease cluster in males and the gout, anaemia, and kidney disease cluster in females, with adjusted HRs of 1.63 (95% CI 1.47–1.81) and 1.96 (95% CI 1.74–2.21), respectively. Among females, the metabolic cluster was associated with a modestly higher mortality risk (aHR 1.20, 95% CI 1.06–1.36). The depressive symptoms, osteoporosis, and frailty-related cluster was also associated with higher mortality risk in both sexes, with aHRs of 1.56 (95% CI 1.40–1.73) in males and 1.68 (95% CI 1.49–1.89) in females.

**Figure 3.**
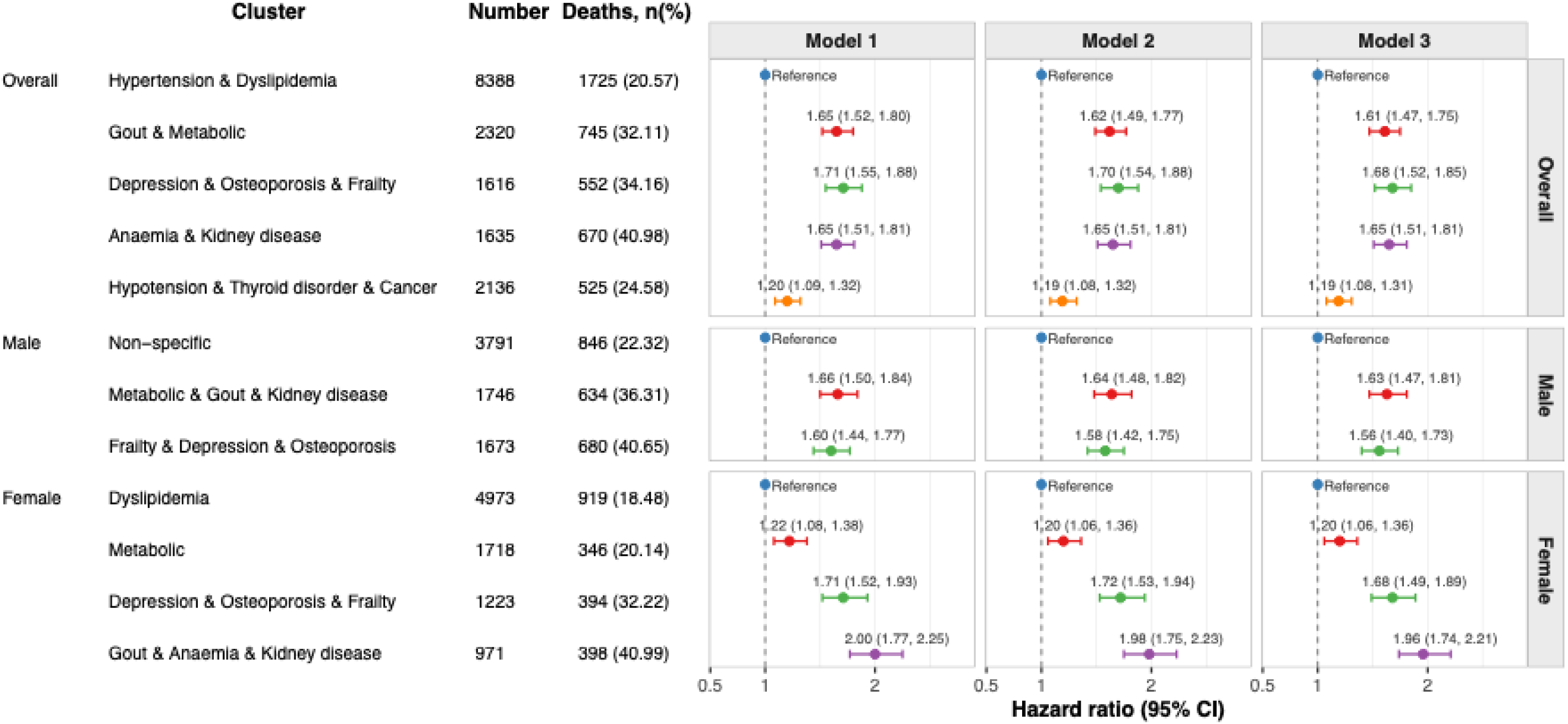
Associations between baseline multimorbidity clusters and all-cause mortality overall and by sex. “Metabolic” in cluster labels indicates obesity and diabetes. Model 1 was adjusted for age and sex in the overall sample and for age only in sex-specific analyses. Model 2 was additionally adjusted for education, area of residence, and living arrangement. Model 3 was further adjusted for smoking status and alcohol consumption. Alt text: Forest plots showing hazard ratios and 95% confidence intervals for associations between baseline multimorbidity clusters and all-cause mortality across three adjustment models in the overall sample, males, and females.

Sensitivity analyses showed broadly similar results to the main analysis. After additional adjustment for the individual-level number of chronic conditions, associations were generally attenuated but remained consistent in the overall sample, and remained significant for all sex-specific clusters except the metabolic cluster in females **(eTable 6).** Analyses using 5-year all-cause mortality showed similar trends in both the overall sample and by sex, with wider confidence intervals due to the smaller number of events **(eFigure 1).**

## Discussion

In this large cohort of relatively healthy community-dwelling older adults aged 70 years and over, we applied LCA to identify distinct multimorbidity profiles based on similar clusters of co-occurring conditions and examined their associations with mortality(8,23). We found that multimorbidity was highly prevalent and clusters differed by sex and in their association with all-cause mortality. These findings suggest that sex-specific clustering of chronic conditions may provide additional prognostic insight beyond the presence or number of individual chronic conditions.

A key difference between our study and many previous multimorbidity studies is that the ASPREE participants were without major cardiovascular disease events at enrolment, and thus our clusters only included chronic conditions rather than major clinical vascular diseases, such as stroke, coronary artery disease, heart failure, and peripheral artery disease(24). Importantly, previous studies have already described cardiovascular-dominant multimorbidity clusters and their strong associations with mortality^7–9^. However, when established cardiovascular diseases are included in clustering analyses, they may strongly influence cluster formation and potentially mask more subtle clusters involving other chronic conditions(28,29). By excluding participants with major cardiovascular disease at baseline, our study was less likely to be dominated by cardiovascular conditions, allowing identification of more diverse multimorbidity clusters and providing insight into disease clustering in otherwise healthy older adults.

The dominant cluster themes identified in the overall population were broadly consistent with previously reported multimorbidity profiles, including metabolic, mental health, and physical function-related clusters(8,12,23,28,30,31). In our study, five clusters were identified, differing in the distribution of chronic conditions across individuals, and sex-stratified analyses revealed that cluster composition varied between males and females. Consistent with previous studies, clusters characterized by hypertension and dyslipidemia, as well as by diabetes and obesity conditions, were observed(8,29). The depressive symptoms, osteoporosis, and frailty-related cluster appeared in both the overall population and in each sex, although the relative representation of component conditions differed between males and females. Because the LCA was conducted separately for each sex, these differences reflect variations in the latent multimorbidity structure rather than merely differences in the distribution of the overall clusters.

Additionally, a gout-and kidney disease-related cluster, which also included an over-representation of diabetes, was observed in both sexes; in males, it also clustered with obesity, while in females with anaemia. This may reflect sex-specific differences in uric acid metabolism, cardiometabolic risk, alcohol consumption, smoking, physical activity, and BMI(32).

Distinct multimorbidity clusters exhibited different mortality risks, with notable sex-specific clusters. In analyses of the overall population, three clusters showed similarly higher mortality risk compared with the least distinctive cluster, whereas clusters characterized by hypotension, thyroid disorders, or cancer history exhibited relatively lower risk. This may reflect the generally stable or well-controlled nature of these conditions in this relatively healthy older population, and the fact that participants with a history of cancer or thyroid disorders also appear in other clusters, which could dilute their apparent contribution to risk(33,34). Sex-stratified analyses revealed more nuanced patterns. In males, two clusters had comparable mortality, although the metabolic, gout-, and kidney disease-related cluster showed a higher risk. In females, the metabolic cluster showed a weaker association with mortality. This may reflect the complex relationship between BMI and mortality in older adults, in which apparent protective effects of higher BMI can arise from reverse causation, illness-related weight loss, selective survival, or differences in body composition rather than a true causal effect(35). Even for the depressive symptoms, osteoporosis, and frailty-related cluster, which was associated with higher mortality in both sexes(30), factors such as depression prevalence, osteoporosis risk, age, hormonal changes, medication use, and physical vulnerability may shape how these conditions co-occur and influence mortality(36–39). Overall, these findings highlight the need for sex-stratified analyses, as pooled analyses may conceal cluster–outcome associations that differ between men and women(7). Such differences arise from sex-specific variation in the organisation of multimorbidity and in biological, behavioural, healthcare-related, and survival-related factors(25,32,40,41).

The co-occurrence of chronic conditions observed in our study is likely to reflect structured and clinically meaningful clusters rather than random aggregation. Similar to metabolic syndrome, in which related metabolic abnormalities cluster together and jointly increase cardiovascular risk(42,43), multimorbidity clusters may represent shared underlying pathways and cumulative physiological dysregulation. The clustering of kidney disease with anaemia may be explained by the established association between renal dysfunction and anaemia(8,44,45), as impaired kidney function can contribute to anaemia through reduced erythropoietin production, inflammation, and disturbances in iron metabolism(46,47). The depressive symptoms, osteoporosis, and frailty-related cluster may similarly represent the convergence of mental health burden and physical vulnerability. Previous evidence suggests that depression in combination with somatic conditions is associated with worse health outcomes than either condition alone, which may help explain the elevated mortality observed for this cluster(37). Given its consistent appearance and association with higher mortality, this pattern warrants particular attention for potential clinical screening and intervention strategies. The hypotension-, thyroid disorder-, and past cancer-related cluster differed from commonly reported clusters. Although hypotension is less commonly included in clustering studies, in older adults, it may indicate complex health status, impaired haemodynamic regulation, medication effects, or underlying disease burden, while thyroid disorder and cancer history may add further clinical complexity(48–50). Taken together, this cluster may represent a distinctive, although heterogeneous, vulnerability profile(51).

Beyond their prognostic value, multimorbidity clusters may have important implications for research and clinical care. Identifying frequently co-occurring conditions can provide a basis for investigating shared disease mechanisms, common risk pathways, and the natural history of multimorbidity(29). Clinically, attention is often directed towards high-priority diseases or conditions, which may lead to other coexisting conditions being overlooked or undertreated. Cluster information may help by identifying over-represented or readily detectable sentinel conditions that prompt screening for other commonly coexisting conditions within the same multimorbidity profile(23,25). This approach also supports a shift from disease-centred care to person-centred management, where treatment goals, medication burden, functional status, and patient priorities are considered within the broader multimorbidity profile(10,23). In the longer term, stable and clinically meaningful clusters may inform guidelines, intervention strategies, and clinical trial designs that better reflect the complexity of older adults living with multiple chronic conditions(25). The identification of sex-specific clusters in our study further suggests that such approaches should account for differences in multimorbidity profiles between males and females.

This study has several strengths. The large, community-dwelling ASPREE cohort with long follow-up enabled robust examination of multimorbidity clusters and mortality in adults aged 70 years and over. By excluding participants with major cardiovascular disease at baseline, clusters were less dominated by vascular conditions, allowing clearer characterisation of other chronic disease profiles. The use of LCA and sex-stratified analyses provided data-driven, clinically interpretable clusters and identified sex-specific multimorbidity structures, enhancing understanding of multimorbidity phenotypes and mortality risk in older adults.

Several limitations should be noted. First, the trial-based selection limits generalisability, particularly to older adults with established cardiovascular disease, dementia, or those in institutional care. Second, only 15 chronic conditions were included, so clusters may differ with a broader or alternative set. Third, LCA does not account for disease severity, which could affect cluster interpretation and mortality risk. Finally, multimorbidity was assessed only at baseline, with subsequent incident conditions, disease progression, and treatment changes not captured, potentially influencing observed associations.

In conclusion, distinct multimorbidity clusters were identified among relatively healthy older adults, with cluster profiles differing between males and females. Several clusters, particularly those characterised by frailty, depressive symptoms, osteoporosis, and kidney disease, were associated with higher all-cause mortality. These findings suggest that specific patterns of disease co-occurrence may provide prognostic information beyond multimorbidity alone and support more tailored approaches to risk stratification and management in older adults.

## Supporting information

eTable 1, eTable 2, eTable 3, eTable 4, eTable 5, eTable 6, eFigure 1

## Data Availability

All data produced in the present work are contained in the manuscript.

## Funding

This work was supported by the National Institute on Ageing and the National Cancer Institute at the National Institutes of Health (U01AG029824 and U19AG062682); the National Health and Medical Research Council (NHMRC) of Australia (334047 and 1127060); Monash University (Australia) and the Victorian Cancer Agency (Australia). Rong Du is supported by an Australian Government Research Training Program (RTP) Scholarship and a Monash International Tuition Scholarship (MITS). Aung Zaw Zaw Phyo is supported by the National Heart Foundation of Australia—Postdoctoral Fellowship (110205-2025_PDF). Joanne Ryan is supported by a National Health and Medical Research Council Research Leadership Investigator Grant (2016438). The sponsors were not involved in the design, methods, subject recruitment, data collection, analysis, or preparation of the paper.

## Acknowledgements

The authors sincerely thank the ASPREE and ASPREE-XT participants for their valuable time and commitment. The authors also gratefully acknowledge the contributions of the study teams in Australia and the United States, participating general practitioners, medical clinics, and supporting organisations involved in the conduct of ASPREE and ASPREE-XT. During manuscript preparation, AI-assisted technology (ChatGPT) was used to assist with grammar and spelling checks. All content was reviewed and approved by the authors.

## Conflict of Interest

Kerry Sheets reports grants from the American Association of Retired Persons and Optum Labs, National Institute on Ageing, and Agency for Healthcare Research and Quality, as well as honoraria from the International Antiviral Society USA. Joanne Ryan reports funding from the National Institute of Health, the National Health and Medical Research Council, the Medical Research Future Fund, and the Australian Research Council, honoraria from the Victorian Coronial Council–Department of Justice and Community Service, and serving as a non-paid committee member and a scientific board member for Dementia Australia. No other competing interests were reported.

## Supplemental Material Legend

eTable 1: Definitions of baseline chronic conditions.

eTables 2–4: Baseline characteristics of participants overall and by sex-specific multimorbidity clusters.

eTable 5: Incidence rates of all-cause mortality across multimorbidity clusters.

eTable 6: Sensitivity analysis of associations between multimorbidity clusters and all-cause mortality with additional adjustment for the number of chronic conditions.

eFigure 1. Associations between baseline multimorbidity clusters and 5-year all-cause mortality in the overall sample and by sex. 7

## Notes

### Author Declarations

ASPREE was approved in Australia by the Royal Australian College of General Practitioners Ethics Committee, the Monash University Human Research Ethics Committee, the Human Research Ethics Committee Tasmanian Network, the Goulburn Valley Health Ethics and Research Committee, the ACT Health Human Research Ethics Committee, and the University of Adelaide Human Research Ethics Committee. In the United States, individual clinic sites obtained Institutional Review Board approval from their respective institutions. ASPREE-XT was overseen in Australia by the Alfred Health Human Research Ethics Committee as the primary ethics site, with approval also from the University of Tasmania Human Research Ethics Committee Network and acceptance by relevant Human Research Ethics Committees through the National Mutual Acceptance Scheme. In the United States, ASPREE-XT used a Single Institutional Review Board of record under the responsibility of the University of Iowa, with a separate Institutional Review Board for the Veterans Affairs site at Emory, Georgia. All participants provided written informed consent.

