## Supplementary material for "Sex-specific multimorbidity clusters and all-cause mortality in relatively healthy older adults: findings from the ASPREE cohort": eTable 1, eTable 2, eTable 3, eTable 4, eTable 5, eTable 6, eFigure 1

**Supplementary tables and figures**

### eTable 1. Definitions of the 15 baseline chronic conditions included in the analysis

| Condition | Definition |
|---|---|
| Hypertension | Mean of three seated blood pressure measurements with a mean systolic blood pressure (SBP) ≥ 140 mmHg, or mean diastolic blood pressure (DBP) ≥ 90 mmHg, or current use of antihypertensive medication. |
| Hypotension | Mean of three seated blood pressure measurements with a mean SBP < 90 mmHg or mean DBP < 60 mmHg. |
| Diabetes | Self-reported diabetes, fasting blood glucose > 125 mg/dL, or current pharmaceutical treatment for diabetes. |
| Dyslipidemia | Use of cholesterol-lowering medication, serum total cholesterol ≥ 212 mg/dL, or low-density lipoprotein cholesterol > 160 mg/dL (> 4.1 mmol/L). |
| Obesity | Body mass index (BMI) ≥ 30 kg/m². |
| Thyroid disorder | Current use of medication for thyroid disorder. |
| Obstructive pulmonary disease | Self-reported asthma or current use of medication for obstructive pulmonary disease. |
| Kidney disease | Self-reported kidney disease, estimated glomerular filtration rate (eGFR) <60 mL/min/1.73 m², or spot urine albumin-to-creatinine ratio (ACR) ≥3 mg/mmol. |
| Pre-Frailty/Frailty | Defined as a Fried frailty phenotype score ≥1, including both pre-frail and frail participants. |
| Cancer history | Self-reported history of cancer. |
| Bowel polyp | Self-reported history of bowel polyp. |
| Gout | Self-reported gout or current use of medication for gout. |
| Osteoporosis | Use of osteoporosis-related medication. |
| Anaemia | Haemoglobin < 13 g/dL in men and < 12 g/dL in women. |
| Depressive symptoms | Center for Epidemiologic Studies Depression Scale 10-item version (CESD-10) score ≥ 8. |

**eTable 2. Baseline characteristics of participants according to multimorbidity clusters in the overall sample**

| Characteristics | Hypertension & Dyslipidemia N = 8,388 | Gout & Metabolic N = 2,320 | Depression, Osteoporosis & Frailty N = 1,616 | Anaemia & Kidney disease N = 1,635 | Hypotension, Thyroid disorder & Cancer N = 2,136 | P value |
|---|---|---|---|---|---|---|
| Age, mean (SD) | 74.71 (3.94) | 75.34 (4.22) | 76.08 (4.56) | 77.89 (5.30) | 74.97 (4.30) | <0.001 |
| Sex, n (%) | | | | | | <0.001 |
| Female | 4523 (53.92) | 1257 (54.18) | 1117 (69.12) | 835 (51.07) | 1153 (53.98) | |
| Male | 3865 (46.08) | 1063 (45.82) | 499 (30.88) | 800 (48.93) | 983 (46.02) | |
| Age group, n (%) | | | | | | <0.001 |
| 70–74 | 5301 (63.20) | 1294 (55.78) | 802 (49.63) | 600 (36.70) | 1321 (61.84) | |
| 75–79 | 2104 (25.08) | 678 (29.22) | 490 (30.32) | 485 (29.66) | 517 (24.20) | |
| 80+ | 983 (11.72) | 348 (15.00) | 324 (20.05) | 550 (33.64) | 298 (13.95) | |
| Education, n (%) | | | | | | <0.001 |
| ≥12 years | 4344 (51.79) | 969 (41.77) | 764 (47.28) | 787 (48.13) | 1142 (53.46) | |
| <12 years | 4044 (48.21) | 1351 (58.23) | 852 (52.72) | 848 (51.87) | 994 (46.54) | |
| Residence area, n (%) | | | | | | <0.001 |
| Outer region | 1026 (12.23) | 308 (13.28) | 138 (8.54) | 172 (10.52) | 242 (11.33) | |
| Inner region | 3064 (36.53) | 844 (36.38) | 572 (35.40) | 550 (33.64) | 713 (33.38) | |
| Major cities | 4298 (51.24) | 1168 (50.34) | 906 (56.06) | 913 (55.84) | 1181 (55.29) | |
| Living status, n (%) | | | | | | <0.001 |
| Living with someone | 5980 (71.29) | 1525 (65.73) | 970 (60.02) | 1031 (63.06) | 1463 (68.49) | |
| Living alone at home | 2408 (28.71) | 795 (34.27) | 646 (39.98) | 604 (36.94) | 673 (31.51) | |
| Smoking status, n (%) | | | | | | <0.001 |
| Current | 275 (3.28) | 68 (2.93) | 66 (4.08) | 45 (2.75) | 90 (4.21) | |
| Former | 3387 (40.38) | 1047 (45.13) | 679 (42.02) | 697 (42.63) | 772 (36.14) | |
| Never | 4726 (56.34) | 1205 (51.94) | 871 (53.90) | 893 (54.62) | 1274 (59.64) | |
| Alcohol consumption, n (%) | | | | | | <0.001 |
| Current | 6775 (80.77) | 1692 (72.93) | 1238 (76.61) | 1270 (77.68) | 1725 (80.76) | |
| Former | 345 (4.11) | 151 (6.51) | 91 (5.63) | 100 (6.12) | 94 (4.40) | |
| Never | 1268 (15.12) | 477 (20.56) | 287 (17.76) | 265 (16.21) | 317 (14.84) | |
| Chronic condition count, n (%) | | | | | | <0.001 |

| | | | | | | |
|---|---|---|---|---|---|---|
| 0 | 0 (0.00) | 0 (0.00) | 0 (0.00) | 0 (0.00) | 319 (14.93) | |
| 1 | 1197 (14.27) | 0 (0.00) | 0 (0.00) | 0 (0.00) | 374 (17.51) | |
| ≥2 | 7191 (85.73) | 2320 (100.00) | 1616 (100.00) | 1635 (100.00) | 1443 (67.56) | |
| Chronic condition count, mean (SD) | 2.63 (1.02) | 5.08 (1.25) | 4.60 (1.29) | 4.35 (1.07) | 1.98 (1.24) | <0.001 |

Note: "Metabolic" in the labels indicates obesity and diabetes.

**eTable 3. Baseline characteristics of male participants according to multimorbidity clusters**

| Characteristics | Non-specific (N = 3,791) | Metabolic, Gout & Kidney disease (N = 1,746) | Frailty, Depression & Osteoporosis ( N = 1,673) | P value |
|---|---|---|---|---|
| Age, mean (SD) | 74.29 (3.66) | 75.49 (4.44) | 76.64 (5.05) | <0.001 |
| Age group, n (%) | | | | <0.001 |
| 70–74 | 2564 (67.63) | 960 (54.98) | 790 (47.22) | |
| 75–79 | 865 (22.82) | 504 (28.87) | 463 (27.67) | |
| 80+ | 362 (9.55) | 282 (16.15) | 420 (25.10) | |
| Education, n (%) | | | | <0.001 |
| ≥12 years | 2123 (56.00) | 824 (47.19) | 871 (52.06) | |
| <12 years | 1668 (44.00) | 922 (52.81) | 802 (47.94) | |
| Residence area, n (%) | | | | 0.133 |
| Outer region | 434 (11.45) | 206 (11.80) | 191 (11.42) | |
| Inner region | 1408 (37.14) | 617 (35.34) | 563 (33.65) | |
| Major cities | 1949 (51.41) | 923 (52.86) | 919 (54.93) | |
| Living status, n (%) | | | | <0.001 |
| Living with someone | 3152 (83.14) | 1359 (77.84) | 1250 (74.72) | |
| Living alone at home | 639 (16.86) | 387 (22.16) | 423 (25.28) | |
| Smoking status, n (%) | | | | <0.001 |
| Current | 134 (3.53) | 61 (3.49) | 83 (4.96) | |
| Former | 1914 (50.49) | 1023 (58.59) | 898 (53.68) | |
| Never | 1743 (45.98) | 662 (37.92) | 692 (41.36) | |
| Alcohol consumption, n (%) | | | | 0.001 |
| Current | 3294 (86.89) | 1469 (84.14) | 1392 (83.20) | |
| Former | 187 (4.93) | 123 (7.04) | 111 (6.63) | |
| Never | 310 (8.18) | 154 (8.82) | 170 (10.16) | |
| Chronic condition count, n (%) | | | | <0.001 |
| 0 | 204 (5.38) | 0 (0.00) | 0 (0.00) | |
| 1 | 759 (20.02) | 0 (0.00) | 72 (4.30) | |
| ≥2 | 2828 (74.60) | 1746 (100.00) | 1601 (95.70) | |
| Chronic condition count, mean (SD) | 2.22 (1.09) | 4.83 (1.24) | 3.27 (1.20) | <0.001 |

Note: “Metabolic” in the labels indicates obesity and diabetes.

### eTable 4. Baseline characteristics of female participants according to multimorbidity clusters

| Characteristics | Dyslipidemia (N = 4,973) | Metabolic (N = 1,718) | Depression, Osteoporosis & Frailty (N = 1,223) | Gout, Anaemia & Kidney disease ( N = 971) | P value |
|---|---|---|---|---|---|
| Age, mean (SD) | 75.05 (4.14) | 74.56 (3.68) | 76.28 (4.67) | 77.90 (5.28) | <0.001 |
| Age group, n (%) | | | | | <0.001 |
| 70–74 | 2971 (59.74) | 1097 (63.85) | 590 (48.24) | 346 (35.63) | |
| 75–79 | 1311 (26.36) | 456 (26.54) | 370 (30.25) | 305 (31.41) | |
| 80+ | 691 (13.90) | 165 (9.60) | 263 (21.50) | 320 (32.96) | |
| Education, n (%) | | | | | <0.001 |
| ≥12 years | 2480 (49.87) | 748 (43.54) | 571 (46.69) | 389 (40.06) | |
| <12 years | 2493 (50.13) | 970 (56.46) | 652 (53.31) | 582 (59.94) | |
| Residence area, n (%) | | | | | 0.005 |
| Outer region | 599 (12.05) | 227 (13.21) | 111 (9.08) | 118 (12.15) | |
| Inner region | 1748 (35.15) | 635 (36.96) | 421 (34.42) | 351 (36.15) | |
| Major cities | 2626 (52.81) | 856 (49.83) | 691 (56.50) | 502 (51.70) | |
| Living status, n (%) | | | | | <0.001 |
| Living with someone | 3021 (60.75) | 999 (58.15) | 674 (55.11) | 514 (52.94) | |
| Living alone at home | 1952 (39.25) | 719 (41.85) | 549 (44.89) | 457 (47.06) | |
| Smoking status, n (%) | | | | | <0.001 |
| Current | 156 (3.14) | 32 (1.86) | 57 (4.66) | 21 (2.16) | |
| Former | 1491 (29.98) | 558 (32.48) | 407 (33.28) | 291 (29.97) | |
| Never | 3326 (66.88) | 1128 (65.66) | 759 (62.06) | 659 (67.87) | |
| Alcohol consumption, n (%) | | | | | <0.001 |
| Current | 3836 (77.14) | 1178 (68.57) | 900 (73.59) | 631 (64.98) | |
| Former | 169 (3.40) | 82 (4.77) | 63 (5.15) | 46 (4.74) | |
| Never | 968 (19.47) | 458 (26.66) | 260 (21.26) | 294 (30.28) | |
| Chronic condition count, n (%) | | | | | <0.001 |
| 0 | 115 (2.31) | 0 (0.00) | 0 (0.00) | 0 (0.00) | |
| 1 | 740 (14.88) | 0 (0.00) | 0 (0.00) | 0 (0.00) | |
| ≥2 | 4118 (82.81) | 1718 (100.00) | 1223 (100.00) | 971 (100.00) | |
| Chronic condition count, mean (SD) | 2.53 (1.11) | 4.11 (1.11) | 4.47 (1.23) | 5.35 (1.29) | <0.001 |

Note: “Metabolic” in the labels indicates obesity and diabetes.

### eTable 5. Mortality incidence rates across baseline multimorbidity clusters during follow-up

| Multimorbidity cluster | Deaths / person-years | Incidence rate per 1,000 person-years (95% CI) |
|---|---|---|
| **Overall** | **4,217 / 167,030** | **25.25 (24.49–26.02)** |
| Hypertension & Dyslipidemia | 1,725 / 89,348 | 19.31 (18.41–20.24) |
| Gout & Metabolic | 745 / 23,381 | 31.86 (29.62–34.24) |
| Depression, Osteoporosis & Frailty | 552 / 16,111 | 34.26 (31.46–37.24) |
| Anaemia & Kidney disease | 670 / 15,867 | 42.22 (39.09–45.55) |
| Hypotension, Thyroid disorder & Cancer | 525 / 22,323 | 23.52 (21.55–25.62) |
| **Male** | **2,160 / 73,583** | **29.35 (28.13–30.62)** |
| Non-specific | 846 / 40,062 | 21.12 (19.72–22.59) |
| Metabolic, Gout & Kidney disease | 634 / 17,128 | 37.02 (34.19–40.01) |
| Frailty, Depression & Osteoporosis | 680 / 16,393 | 41.48 (38.42–44.72) |
| **Female** | **2,057 / 93,447** | **22.01 (21.07–22.98)** |
| Dyslipidemia | 919 / 53,372 | 17.22 (16.12–18.37) |
| Metabolic | 346 / 18,199 | 19.01 (17.06–21.12) |
| Depression, Osteoporosis & Frailty | 394 / 12,372 | 31.85 (28.78–35.15) |
| Gout, Anaemia & Kidney disease | 398 / 9,504 | 41.88 (37.86–46.20) |

Note: “Metabolic” in the labels indicates obesity and diabetes.

### eTable 6. Sensitivity analysis of the association between baseline multimorbidity clusters and all-cause mortality after additional adjustment for the individual-level number of chronic conditions

| Multimorbidity cluster | aHR (95% CI) | P value |
|---|---|---|
| **Overall** | | |
| Hypertension & Dyslipidemia | 1.00 (Ref.) | |
| Gout & Metabolic | 1.34 (1.20–1.49) | <0.001 |
| Depression, Osteoporosis & Frailty | 1.46 (1.31–1.63) | <0.001 |
| Anaemia & Kidney disease | 1.46 (1.32–1.61) | <0.001 |
| Hypotension, Thyroid disorder & Cancer | 1.25 (1.13–1.38) | <0.001 |
| **Male** | | |
| Non-specific | 1.00 (Ref.) | |
| Metabolic, Gout & Kidney disease | 1.38 (1.20–1.59) | <0.001 |
| Frailty & Depression & Osteoporosis | 1.46 (1.31–1.63) | <0.001 |
| **Female** | | |
| Dyslipidemia | 1.00 (Ref.) | |
| Metabolic | 1.09 (0.95–1.25) | 0.216 |
| Depression, Osteoporosis & Frailty | 1.50 (1.30–1.71) | <0.001 |
| Gout, Anaemia & Kidney disease | 1.67 (1.43–1.94) | <0.001 |

Note: “Metabolic” in the labels indicates obesity and diabetes. Cox proportional hazards models were adjusted for sex (overall sample) and for age, education, residence area, living status, smoking status, alcohol consumption, and number of chronic conditions.

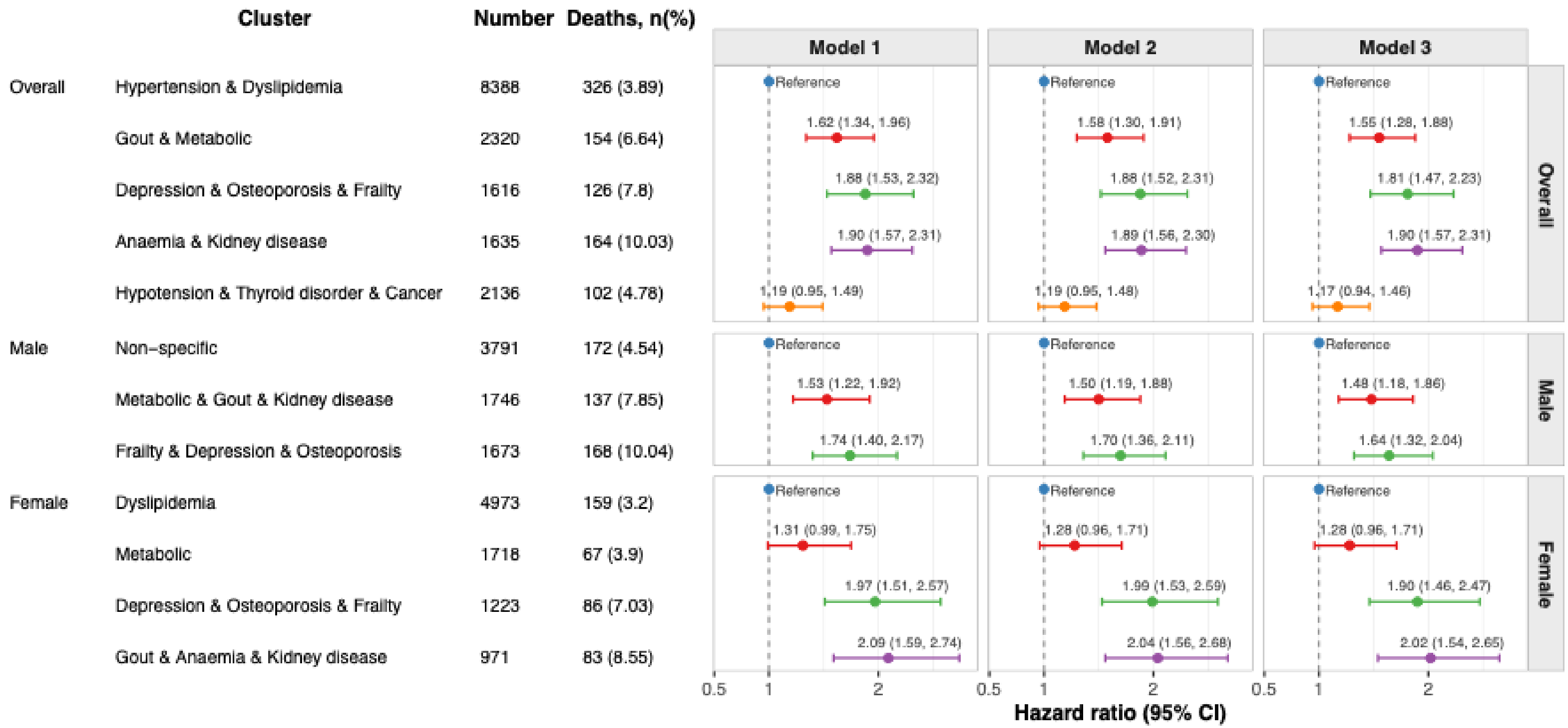

**eFigure 1. Associations between baseline multimorbidity clusters and 5-year all-cause mortality in the overall sample and by sex.**

Note: “Metabolic” in the labels indicates obesity and diabetes. Model 1 was adjusted for age and sex (overall sample) or age only (sex-specific analyses). Model 2 was additionally adjusted for education, residence area, and living status. Model 3 was further adjusted for smoking status and alcohol consumption.
